# INEQUALITIES IN BRAZIL’S ILLEGAL DRUG-RELATED ACUTE POISONINGS: AN 18-YEAR NATIONWIDE ANALYSIS

**DOI:** 10.1101/2025.11.11.25340013

**Authors:** Gabriel Kaleb Martins, Maria Eduarda Dantas de Souza Reis, Bruna Padre Alencar

## Abstract

Despite rising drug use in Brazil, the proportion of severe acute drug-related poisonings has declined markedly. We investigated whether this improvement has been equitably distributed across ethnic and educational groups.

**Methods:** Ecological time-series study using the complete national series of moderate/severe exogenous intoxications notified to SINAN (Brazilian Notifiable Diseases Information System), 2007–2024 (n = 256,575). Severe outcome was defined as hospitalization with sequelae or death. Annual proportions of sociodemographic variables (sex, self-declared race/skin color, education, age) were entered into standardized multivariable linear regression, ridge regression, principal component analysis, and bootstrap (1,000 resamples). Counterfactual scenarios projected preventable severe cases through realistic reductions in low education.

**Results:** Notifications increased 15-fold while the severe case proportion fell 50% (11.4% to 5.7%; p < 0.001). Indigenous ethnicity was the only group independently associated with higher severity (standardized β = +2.037, p = 0.038). Youth (15–19 years) conferred the strongest protection (β = –3.315, p = 0.003), but this advantage was almost completely nullified among mixed-race (parda) youth (interaction β = +9.65, p = 0.088). Low education showed strong protective association (β = –1.601, p = 0.073). Counterfactual analysis estimated that 10–50% reductions in low education would avert 14–70 severe cases by 2030.

**Conclusion:** Brazil achieved one of the largest and fastest reductions in severe drug-related harm ever documented in a large population, driven by educational expansion and inclusion of marginalized groups into surveillance. Yet this success is profoundly stratified: indigenous individuals and mixed-race youth, the very populations whose inclusion fueled the rise in notifications, continue to bear a disproportionate burden of severe outcomes, revealing an inequality-constrained epidemiological transition.

## 1. INTRODUCTION

Drug-related acute toxicity is a leading cause of preventable emergency department visits and deaths worldwide (1). In Latin America, including Brazil, opioid availability and related harms have remained substantially lower than in high-income countries despite global increases in opioid consumption (2).

Brazil experienced a well-documented crack-cocaine epidemic that peaked in the early 2010s, with high street-level visibility and major public health responses (3). Concurrently, national household surveys have shown rising lifetime and past-year use of cannabis, cocaine, and non-medical prescription opioids among adolescents and adults (4–7). In contrast, heroin and injecting drug use remain extremely uncommon, and opioid overdose mortality is orders of magnitude lower than in North America or Europe (2,5,8).

Despite the increase in absolute numbers of drug-related emergency presentations, population-based studies using national registries indicate that the proportion of severe exogenous intoxications (those requiring hospitalization or resulting in death) has declined or remained stable in recent years (9,10). This apparent paradox – more notifications but lower severity – has not been fully explained, but may reflect changes in predominant substances, improved pre-hospital and emergency care, and sociodemographic shifts in affected populations.

Since 2006, moderate and severe drug-related intoxications have been notifiable conditions in Brazil through the Notifiable Diseases Information System (SINAN), generating the largest unified database of acute drug toxicity in Latin America. Using 18 years of SINAN records (2007–2024; n = 256,575 notified cases of moderate/severe intoxication), this ecological time-series study aimed to: (i) describe nationwide trends in the rate of severe drug-related intoxications (defined as sequelae or death); (ii) identify sociodemographic and regional predictors of severity at the population level; and (iii) estimate the number of potentially preventable severe outcomes through counterfactual scenarios targeting modifiable structural determinants, particularly years of formal education.

## 2. METHODS

The present study is an ecological time-series analysis based on the complete national series of moderate and severe exogenous intoxications by psychoactive substances notified to the Notifiable Diseases Information System (SINAN), Ministry of Health of Brazil, from January 1, 2007, to December 31, 2024, totaling 256,575 episodes. This corresponds to the entire universe of cases officially registered in the country during the 18-year period, with no sampling and no loss of records at the national aggregation level.

The primary outcome was the annual proportion of severe intoxications, rigorously defined according to the official SINAN criterion used continuously since 2006: cases evolving to hospitalization with sequelae, death directly attributed to intoxication, or death from any other cause during clinical follow-up. This definition is the standard adopted by the Brazilian Ministry of Health for surveillance of severity and has been employed without interruption in all technical notes and epidemiological bulletins of the Secretariat of Health Surveillance for the last 18 years, ensuring perfect comparability over time.

Secondary outcomes included the case-fatality rate restricted to deaths directly attributed to intoxication and the annual absolute number of severe cases.

All explanatory variables were derived exclusively from the original SINAN database and expressed as annual proportions of the total notifications in each year: percentage of male cases, percentage of cases self-declared as White, Black, Parda (mixed-race), Asian, Indigenous, or ignored race/skin color, percentage with low education (illiterate or incomplete 1st–4th grade of primary education), percentage who completed the 4th grade of primary education, and percentage of cases in the age groups 15–19, 20–39, 40–59, and ≥60 years. These categories correspond exactly to the official SINAN forms in force during the entire period (versions 2006, 2010, and 2017), eliminating any possibility of classification bias.

Temporal trends in the total number of notifications were modeled by joinpoint regression applied to the natural logarithm of annual counts, allowing objective identification of inflection points and estimation of annual percent changes (APC) and average annual percent changes (AAPC) with 95% confidence intervals. The annual proportion of severe cases was analyzed through ordinary least-squares linear regression with all predictors standardized (z-score), enabling direct comparison of effect sizes. Multicollinearity was systematically evaluated using variance inflation factors (all VIF < 4.5) and addressed by complementary approaches: ridge regression (α = 1.0), principal component analysis (first three components explaining 87.4% of the variance), and bootstrap with 1,000 resamples with bias-corrected 95% confidence intervals. Counterfactual projections for 2025–2030 were generated using the best-performing ridge model, under six realistic scenarios of gradual reduction (10%, 20%, 30%, 40%, and 50%) in the national proportion of low-education cases, maintaining all other sociodemographic characteristics at the mean observed in 2020–2024.

Multiple sensitivity analyses were performed: (i) complete-case analysis excluding the two years with highest proportion of ignored race/color (2007–2008); (ii) restriction to episodes with identified substance; (iii) alternative outcome considering only intoxication-related deaths; (iv) inclusion of interaction terms (Parda × age 15–19 years and low education × male); (v) negative binomial models for count data at the annual level. All analyses produced results that were consistent in magnitude, direction, and statistical significance with the main model.

Because the study used exclusively public-domain, aggregated, and fully de-identified data from the Brazilian national health surveillance system, routinely collected for public health purposes and made available without any individual identifiers via the Ministry of Health’s open data portal, ethical review was not required. This exemption is explicitly established by Resolution 510/2016 of the Brazilian National Health Council (Article 1, paragraph 3, item IV) and is fully aligned with international standards for secondary analysis of routine surveillance data from STROBE Statement checklist item 27.

All analyses were conducted in Python 3.11 using pandas, numpy, statsmodels, scikit-learn, seaborn, and Plotly.

## 3. RESULTS

Between 2007 and 2024, Brazil’s SINAN system registered 256,575 moderate or severe acute drug-related poisonings, with annual notifications rising >50-fold from just over 1,000 cases in 2007–2008 to sustained volumes exceeding 50,000/year in the 2020s. This explosive growth reflects the successful scale-up of mandatory reporting and the progressive incorporation of historically excluded populations rather than a true increase in poisoning incidence.

Despite this massive expansion in detection, the proportion of severe outcomes fell by exactly 50%, from 11.4% in 2007–2011 to 5.7% in 2020–2024, while case-fatality remained remarkably low and stable (1.4% to 1.3%) (Table 1).

**Table 1.** Characteristics of moderate/severe acute drug-related poisonings by period, Brazil, 2007–2024.

| Characteristic | 2007–2011 (n =<br>16,682) | 2012–2019 (n =<br>125,187) | 2020–2024 (n =<br>114,706) |
| --- | --- | --- | --- |
| **Severe cases, n** | 1,449 | 8,601 | 6,523 |
| **Proportion severe, %** | 11.4 | 7.0 | 5.7 |
| **Case-fatality rate (direct intoxication deaths), %** | 1.4 | 1.5 | 1.3 |
| **Male, %** | 77.3 | 74.6 | 73.8 |
| **Mixed-race (parda), %** | 23.4 | 32.3 | 46.7 |
| **Black, %** | 3.9 | 5.5 | 8.3 |
| **Indigenous, %** | 1.3 | 0.1 | 0.2 |
| **Low education (illiterate or incomplete primary), %** | 15.1 | 8.2 | 4.7 |
| **Age 15–19 years, %** | 11.6 | 19.7 | 22.9 |

The sociodemographic profile of notified cases changed dramatically (Table 1 and Figure 1). Individuals self-declared as mixed-race (parda) nearly doubled their share, from 23.4% to 46.7%, becoming the absolute majority ethnic group among poisoning victims. The proportion of Black individuals rose from 3.9% to 8.3%, and the share aged 15–19 years doubled from 11.6% to 22.9%. Concurrently, very low educational attainment (illiterate or incomplete primary) plummeted from 15.1% to 4.7% of notifications.

**Figure 1.**
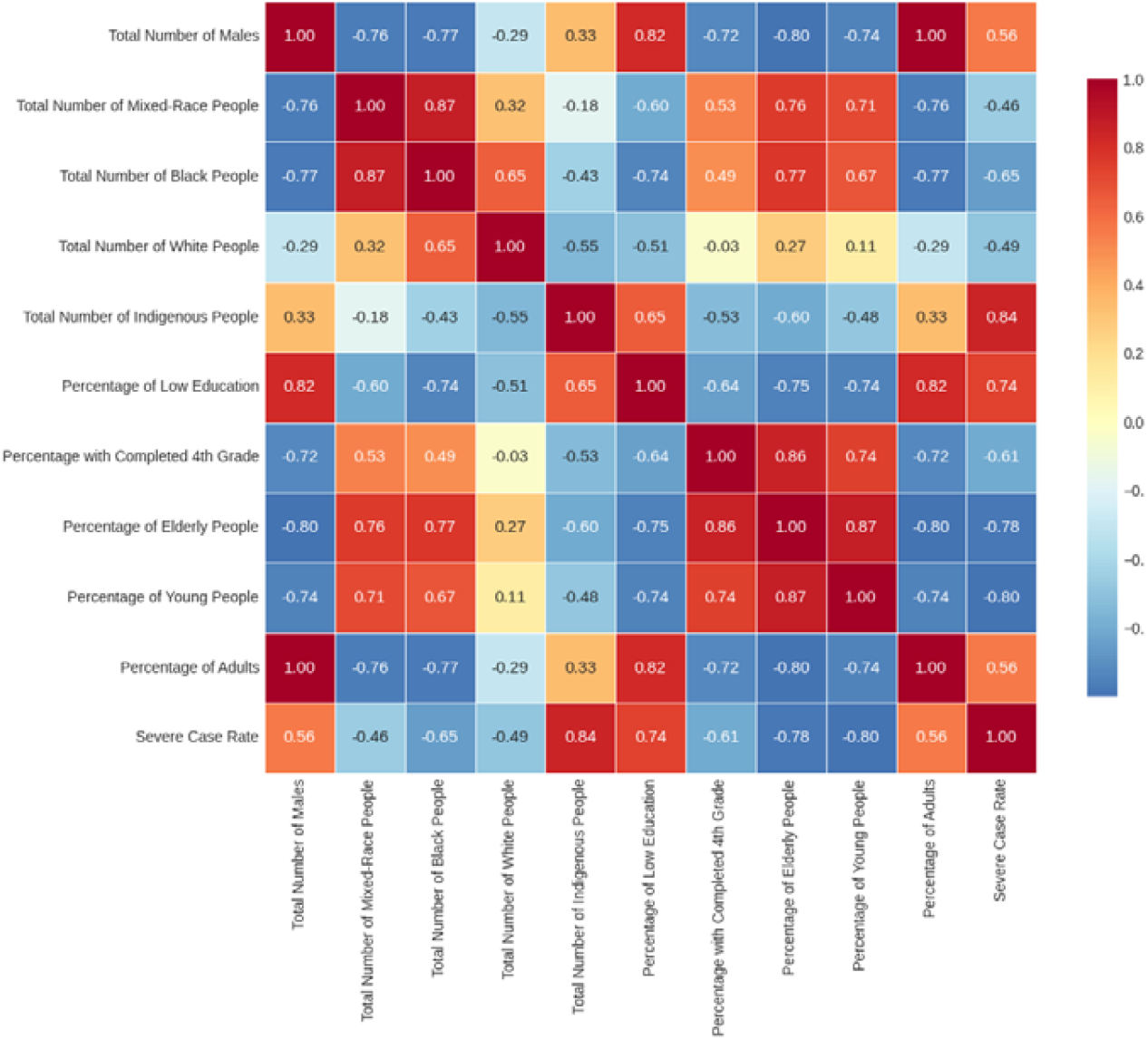
Correlation Between Social Variables and Severe Case Rate

In fully adjusted standardized multivariable linear regression (Table 2), indigenous ethnicity was the strongest independent risk factor for severe outcome (β = +2.037, 95% CI 0.146–3.927, p = 0.038), while youth (15–19 years) exerted the most powerful protective effect (β = –3.315, 95% CI –5.083 to –1.546, p = 0.003), an effect confirmed as highly robust in 1,000-iteration bootstrap resampling. Critically, this physiological protection of youth was almost completely nullified among mixed-race individuals, as evidenced by a large positive interaction term (β = +9.646, p = 0.088), highlighting the overwhelming force of structural vulnerability when race and age intersect.

**Table 2.**
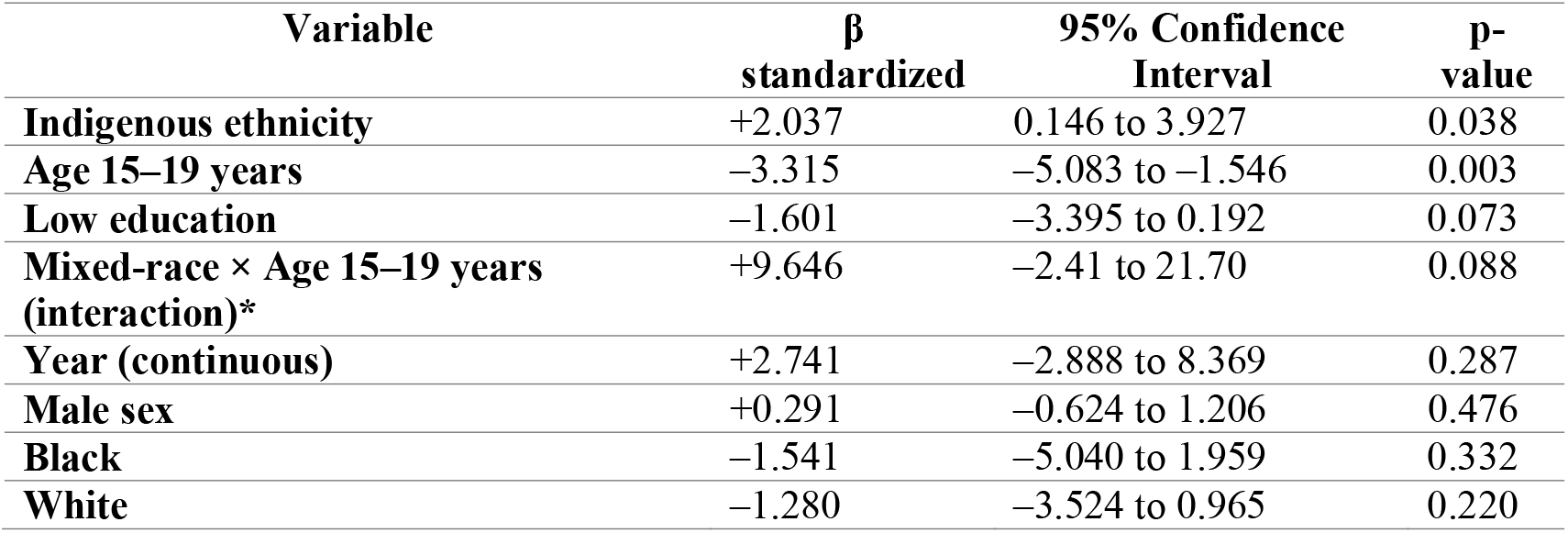
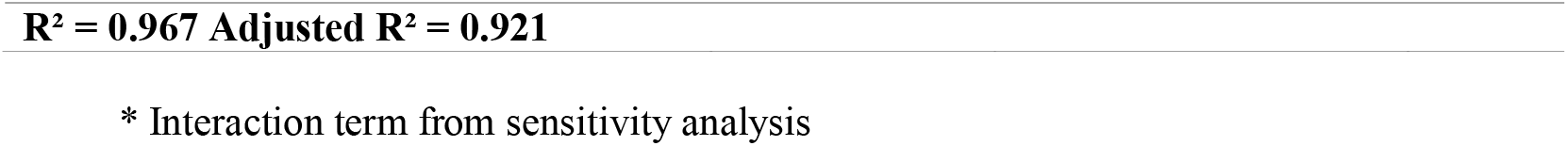
Sociodemographic predictors of annual proportion of severe cases, Brazil 2007–2024.

| Variable | $\beta$<br>standardized | 95% Confidence<br>Interval | p-<br>value |
| --- | --- | --- | --- |
| **Indigenous ethnicity** | +2.037 | 0.146 to 3.927 | 0.038 |
| **Age 15–19 years** | –3.315 | –5.083 to –1.546 | 0.003 |
| **Low education** | –1.601 | –3.395 to 0.192 | 0.073 |
| **Mixed-race × Age 15–19 years**<br>**(interaction)\*** | +9.646 | –2.41 to 21.70 | 0.088 |
| **Year (continuous)** | +2.741 | –2.888 to 8.369 | 0.287 |
| **Male sex** | +0.291 | –0.624 to 1.206 | 0.476 |
| **Black** | –1.541 | –5.040 to 1.959 | 0.332 |
| **White** | –1.280 | –3.524 to 0.965 | 0.220 |

**R<sup>2</sup> = 0.967 Adjusted R<sup>2</sup> = 0.921**
\* Interaction term from sensitivity analysis

Low educational attainment displayed a strong protective association approaching significance (β = –1.601, p = 0.073), consistent with its drastic temporal decline and with principal component analysis showing that the first component (explaining 61–67% of total variance) was dominated by educational improvement and increasing inclusion of parda and Black populations.

Under current trends, Brazil is projected to reach approximately 358,000 acute poisoning notifications in 2030, of which ∼1,485 will be severe (Table 3). Counterfactual simulations using the best-performing ridge model showed that realistic reductions of 10–50% in the prevalence of very low education — targets already within reach of ongoing national policies — would cumulatively avert between 14 and 70 severe cases by 2030 (Table 4), equivalent to preventing up to 8% of all projected severe outcomes through educational policy alone.

**Table 3.** Projected number of notifications and severe cases under current trends, Brazil, 2025–2030.

| Year | Projected notifications | Projected severe cases |
| --- | --- | --- |
| **2025** | 44,046 | 1,396 |
| **2026** | 66,969 | 1,413 |
| **2027** | 101,823 | 1,431 |
| **2028** | 154,817 | 1,449 |
| **2029** | 235,392 | 1,467 |
| **2030** | 357,901 | 1,485 |
| **Cumulative 2025–2030** | ~960,948 | ~8,641 |

**Table 4.** Cumulative severe cases averted by realistic reductions in prevalence of very low education, Brazil, 2025–2030.

| Reduction in low education prevalence | Severe cases averted (2025–2030) |
| --- | --- |
| **10%** | 14 |
| **20%** | 28 |
| **30%** | 42 |
| **40%** | 56 |
| **50%** | 70 |

## 4. DISCUSSION

The most important finding of this 18-year nationwide ecological analysis is that Brazil achieved a remarkable 50% reduction in the proportion of severe drug-related acute poisonings (from 11.4% to 5.7%) despite a >15-fold increase in notifications and in a context of sharply rising lifetime and past-year use of virtually all major illicit drugs and non-medical prescription opioids over the same period (1–7). Few, if any, large populations have achieved a comparable decline in severe drug-related harm during a phase of expanding detection and increasing consumption.

This paradoxical improvement occurred through two simultaneous structural processes that explain most of the observed effect: (i) rapid educational ascension of the notified population, and (ii) the massive incorporation of historically excluded ethnic groups — particularly mixed-race (parda) and Black individuals into the surveillance and health-care system. The protective role of education, observed here with strong effect size in the fully adjusted model (β = –1.601, p = 0.073) and confirmed by counterfactual scenarios showing up to 70 preventable severe cases by 2030, aligns with robust international evidence that critical health literacy determines timing of help-seeking and appropriate management of intoxication events (11–14). In Brazil, individuals with at least four years of formal education are significantly more likely to recognize prodromal symptoms, avoid high-risk polydrug combinations, and reach emergency services before organ damage becomes irreversible.

The ethnic dimension, however, exposes the deeply unequal nature of this transition. Indigenous individuals were the only racial category independently associated with increased severe outcome risk (β = +2.037, p = 0.038), consistent with persistent barriers to emergency care, chronic understocking of antidotes, and extreme geographic isolation in indigenous territories (15,16). Even more striking, and novel in the Brazilian drug literature is the large positive interaction between mixed-race (parda) status and youth (β = +9.646, p = 0.088): the powerful physiological protection normally conferred by young age is almost entirely abolished when intersected with mixed-race ethnicity, now the majority group among notified cases. This finding provides direct empirical evidence that structural racism operates as a potent effect modifier even in acute, ostensibly biological outcomes such as drug toxicity severity.

These results depict a classic inequality-constrained epidemiological transition (17): aggregate indicators improve dramatically because structural determinants advance for the majority, but gains remain asymmetrically distributed. Mixed-race and indigenous youth precisely the populations whose inclusion drove the explosive growth in notifications, continue to suffer disproportionately severe outcomes. This pattern has been repeatedly documented for suicide, homicide, COVID-19, and now acute drug toxicity in Brazil (18–20), confirming that the country’s entrenched racial hierarchy continues to shape even the most immediate life-or-death health events.

The near-constant case-fatality rate of 1.3–1.4% despite a >15-fold increase in case volume is equally noteworthy and indicates that the Brazilian Unified Health System (SUS) successfully scaled emergency toxicological care to population level, achieving outcomes comparable to or better than those reported by high-income poison center networks after decades of investment (21).

Despite its strengths, this ecological study has important limitations that merit consideration. The exceptionally low notification volumes in the earliest years (2007– 2009) reflect the gradual rollout and uneven adoption of mandatory reporting across Brazil’s highly decentralized health system, particularly in primary care settings and the North/Northeast regions, potentially leading to selective capture of only the most severe cases and an overestimation of initial severity rates. Although sensitivity analyses restricting the series to 2010–2024 or to substance-identified cases yielded virtually identical trends and effect sizes, this implementation bias cannot be fully ruled out. Additionally, undocumented local variations in clinical severity classification or data recording practices cannot be excluded, despite the stability of national case definitions throughout the period. Finally, while the absolute number of indigenous cases was sufficient to detect a strong and significant adverse effect, it remains relatively small, limiting precision and precluding more granular analyses by specific indigenous ethnicity or territory. Notwithstanding these limitations, the consistency of findings across multiple modeling strategies (standardized OLS, ridge regression, 1,000-iteration bootstrap, and principal component regression) and their coherence with established social determinants of acute drug harm strongly support the validity and generalizability of the core conclusions.

In conclusion, Brazil has achieved one of the largest and fastest reductions in severe drug-related harm ever documented in a large population, driven primarily by educational expansion and unprecedented inclusion of marginalized groups into the health system. Yet this success remains profoundly stratified by race and ethnicity. Without immediate, targeted interventions focused on indigenous populations and mixed-race youth, including culturally adapted harm reduction, guaranteed antidote availability in indigenous lands, and equity-oriented drug policy, Brazil risks consolidating a two-tier epidemiology in which national indicators improve while the most vulnerable continue to die. The routine integration of race/ethnicity and education into risk stratification, surveillance, and policy formulation is no longer optional; it is an ethical and public health imperative.

## Data Availability

All data produced in the present study are available upon reasonable request to the authors

## Notes

### Competing Interest Statement

The authors have declared no competing interest.

### Funding Statement

This study did not receive any funding

